# Prospective analytical performance evaluation of the QuickNavi™-COVID19 Ag for asymptomatic individuals

**DOI:** 10.1101/2021.04.01.21254813

**Authors:** Yoshihiko Kiyasu, Yuto Takeuchi, Yusaku Akashi, Daisuke Kato, Miwa Kuwahara, Shino Muramatsu, Shigeyuki Notake, Atsuo Ueda, Koji Nakamura, Hiroichi Ishikawa, Hiromichi Suzuki

## Abstract

**Introduction:** Antigen testing may help screen for and detect severe acute respiratory syndrome coronavirus 2 (SARS-CoV-2) infections in asymptomatic individuals. However, limited data regarding the diagnostic performance of antigen tests for this group are available.

**Methods:** We used clinical samples to prospectively evaluate the analytical and clinical performance of the antigen test QuickNavi™-COVID19 Ag. This study was conducted at a PCR center between October 7, 2020 and January 9, 2021. Two nasopharyngeal samples per patient were obtained with flocked swabs; one was used for the antigen test, and the other for real-time reverse transcription PCR (RT-PCR). The diagnostic performance of the antigen test was compared between asymptomatic and symptomatic patients, and the RT-PCR results were used as a reference.

**Results:** Among the 1,934 collected samples, SARS-CoV-2 was detected by real-time RT-PCR in 188 (9.7%); 76 (40.4%) of these samples were from asymptomatic individuals. Over half of the total samples (1,073; 55.5%) were obtained from asymptomatic volunteers. The sensitivity of the antigen test was significantly lower for asymptomatic group than for symptomatic patients (67.1% vs 89.3%, *p* < 0.001). The specificity was 100% for both groups, and no false positives were observed among all 1,934 samples. The median Ct value for the asymptomatic group was significantly higher than that of the symptomatic group (24 vs 20, *p* < 0.001).

**Conclusions:** The QuickNavi™-COVID19 Ag showed a lower sensitivity for asymptomatic group than for symptomatic patients. However, its specificity was consistently high, and no false positives were found in this study.

## Introduction

Severe acute respiratory syndrome coronavirus 2 (SARS-CoV-2), which causes coronavirus disease 2019 (COVID-19), has overwhelmed healthcare systems globally [1]. The early identification and isolation of patients infected with SARS-CoV-2 are essential for constraining COVID-19 transmission.

Travel restrictions have been enforced worldwide to impede the spread of SARS-CoV-2 [2], and many countries have implemented immigration screening measures to minimize the risk of travelers bringing the virus into the country with them [3]; however, the need for resuming domestic and international movement is growing. According to recent data, symptom-based screening, including body-temperature screening, fails to detect a substantial number of SARS-CoV-2-infected patients who have no or mild symptoms [4]. Thus, more accurate screening methods, ideally ones that are convenient and provide rapid results, are desired to detect such individuals.

Although nucleic acid amplification tests (NAATs) are considered highly reliable for detecting SARS-CoV-2, the disadvantages of their finite availability, long turnaround time, and requirement for skilled technicians to perform them have limited their utility for screening purposes [5]. Antigen tests are more accessible point-of-care tests, and they generally take less than an hour to produce results. They can therefore be more beneficial for use in SARS-CoV-2 screening, if their diagnostic performance is sufficient. However, data on the performance of antigen tests in asymptomatic individuals is currently scarce.

Our previous study demonstrated that the antigen test QuickNavi™-COVID19 Ag (Denka Co., Ltd., Tokyo, Japan) had good performance in the detection of patients with COVID-19, with a sensitivity of 86.7% (95% CI: 78.6%–92.5%) and specificity of 100% (95% CI: 99.5%–100%) in 1,186 patients [6]. However, only a few asymptomatic subjects were included in that study, so the diagnostic performance of the QuickNavi™-COVID19 Ag in asymptomatic individuals could not be thoroughly evaluated.

In the present prospective study, we aimed to evaluate the analytical and clinical performance of the QuickNavi™-COVID19 Ag in asymptomatic individuals. This study was conducted as an extension study of our previous report [6].

## Patients and Methods

The details of our study protocol were described previously [6]. Briefly, we prospectively performed this study between October 7, 2020 and January 9, 2021. Sample collection was performed at the PCR center in Tsukuba Medical Center Hospital (TMCH). The enrolled subjects included patients who had been referred from a local public health center or one of 97 primary care facilities and TMCH healthcare workers; clinical information was obtained from each volunteer. All samples from the same patients collected at different timepoints were included in the analysis. The ethics committee of TMCH approved the present study (approval number: 2020-033).

### Sample collection and antigen test procedure

For sample collection, we simultaneously obtained two nasopharyngeal samples: one sample for use in the antigen test, and the other for use in a PCR examination. The antigen test was performed using the QuickNavi™-COVID19 Ag in accordance with the manufacturers’ instructions. The other swab sample was transferred to an in-house microbiology laboratory within an hour of sample collection.

### PCR examinations for SARS-CoV-2

Purification and RNA extraction was performed with magLEAD 6gC (Precision System Science Co., Ltd., Chiba, Japan) for in-house reverse transcription PCR (RT-PCR). The reference real-time RT-PCR of SARS-CoV-2 was performed by using a method developed by the National Institute of Infectious Diseases, Japan [7]. Samples with discrepant results between the reference real-time RT-PCR and in-house RT-PCR were re-evaluated with a GeneXpert^®^ for SARS-CoV-2 (Cepheid Inc., Sunnyvale, CA, USA). The final judgment for the detection of SARS-CoV-2 was based on the GeneXpert^®^ for SARS-CoV-2 result.

### Statistical analyses

The sensitivity and specificity of the QuickNavi™-COVID19 Ag were calculated using the Clopper and Pearson method, with 95% confidence intervals (CIs). Ct values were compared between groups using Mann-Whitney U tests, and *p*-values of <0.05 were considered to indicate statistically significant differences. Categorical variables were compared by using a Fisher’s exact test. All calculations were conducted using the R 4.0.3 software program (www.r-project.org).

## Results

During the study period, we evaluated 1,939 nasopharyngeal samples taken from 1,881 volunteers. After excluding the samples collected from subjects for whom symptom data was unavailable (n = 5), we were left with 1,934 samples for analysis. Of these 1,934 samples, 1,073 (55.5%) were from asymptomatic individuals.

SARS-CoV-2 was detected by reference real-time RT-PCR in 187 samples. There was one discordant sample that produced positive results from the in-house RT-PCR assay but negative results from the reference real-time RT-PCR. This sample was deemed positive for SARS-CoV-2 following an additional examination using GeneXpert for SARS-CoV-2 (Ct value of the N2 gene: 42.6). Thus, 188 of 1,934 total samples (9.7%) were assessed as positive for SARS-CoV-2. Of the 188 SARS-CoV-2-positive samples, 76 (40.4%) were from asymptomatic individuals.

### Sensitivity and specificity of the antigen test in asymptomatic individuals

A comparison of the QuickNavi™-COVID19 Ag assay results with the reference real-time RT-PCR results for samples from asymptomatic volunteers is summarized in Table 1. The sensitivity and specificity were 67.1% (95% CI: 55.4%–77.5%) and 100% (95% CI: 99.4%–100.0%), respectively. Of the 25 samples with discordant results between these two assays, all were assessed as negative by the QuickNavi™-COVID19 Ag assay and assessed as positive by the reference real-time RT-PCR assay (Table 1).

**Table 1.** Sensitivity and specificity of QuickNavi™-COVID19 Ag among asymptomatic individuals.

| Asymptomatic subjects |  | Real-time RT-PCR |  |
| --- | --- | --- | --- |
|  |  | Positive | Negative |
| QuickNavi™-COVID19 Ag | Positive | 51 | 0 |
|  | Negative | 25 | 997 |
| Sensitivity (%) |  | 67.1 (55.4–77.5) |  |
| Specificity (%) |  | 100 (99.4–100) |  |
| Positive predictive value (%) |  | 100 (89.7–100) |  |
| Negative predictive value (%) |  | 97.6 (96.4–98.4) |  |

RT-PCR, reverse transcription-polymerase chain reaction
Data in parentheses are 95% confidence intervals.

The overall sensitivity and specificity of the antigen test were 80.3% (95% CI: 73.9%– 85.7%) and 100% (95% CI: 99.7%–100%), respectively, and no false-positive results were identified among the 1,934 samples. The sensitivity of the QuickNavi™-COVID19 Ag was 89.3 % (95% CI: 82.0%–94.3%) for symptomatic patients, which is significantly higher than its sensitivity for asymptomatic individuals (*p* < 0.001). A comparison of test performance between asymptomatic and symptomatic subjects is summarized in Table 2. Additionally, the QuickNavi™-COVID19 Ag sensitivities stratified by symptom occurrence and Ct value are shown in Table 3.

**Table 2.** Comparison of QuickNavi™-COVID19 Ag performance between asymptomatic and symptomatic subjects.

|  | Asymptomatic subjects<br>(N = 76) | Symptomatic subjects<br>(N = 112) | *p*-value |
| --- | --- | --- | --- |
| Sensitivity (%) | 67.1 (55.4–77.5) | 89.3 (82.0–94.3) | <0.001 |
| Specificity (%) | 100 (99.4–100) | 100 (99.3–100) | 1.000 |
| Positive predictive value (%) | 100 (89.7–100) | 100 (94.6–100) | 1.000 |
| Negative predictive value (%) | 97.6 (96.4–98.4) | 98.4 (97.3–99.2) | 0.919 |

Reverse transcription-polymerase chain reaction was used as the reference.
Data in parentheses are 95% confidence intervals.

**Table 3.** Antigen test sensitivities stratified by Ct values.

| Ct value (N2 gene) | Sensitivity |  |  |
| --- | --- | --- | --- |
|  | Total | Asymptomatic | Symptomatic |
|  | (n = 188) | (n = 76) | (n = 112) |
| Lower than 20 | 100 (91.7–100) | 100 (71.3–100) | 100 (89.1–100) |
| Between 20–25 | 98.2 (90.6–100) | 100 (78.1–100) | 97.1 (85.1–99.9) |
| Between 25–30 | 86.2 (68.3–96.1) | 90.9 (58.7–99.8) | 83.3 (58.6–96.4) |
| 30 or higher | 16.2 (6.2–32.0) | 11.5 (2.4–30.2) | 27.3 (6.0–61.0) |

Data in parentheses are 95% confidence intervals.
Ct, cycle threshold

### Detailed data of samples with discrepant results between antigen test and real-time RT-PCR examinations

Among the 37 samples with discrepant results between the antigen test and real-time RT-PCR examinations, 25 were collected from asymptomatic subjects. The N2 gene was detected in all discrepant samples except one from an asymptomatic individual, which yielded positive results from a subsequent GeneXpert for SARS-CoV-2 assay. The N1 gene was not detected in 18 samples (Table 4).

**Table 4.**
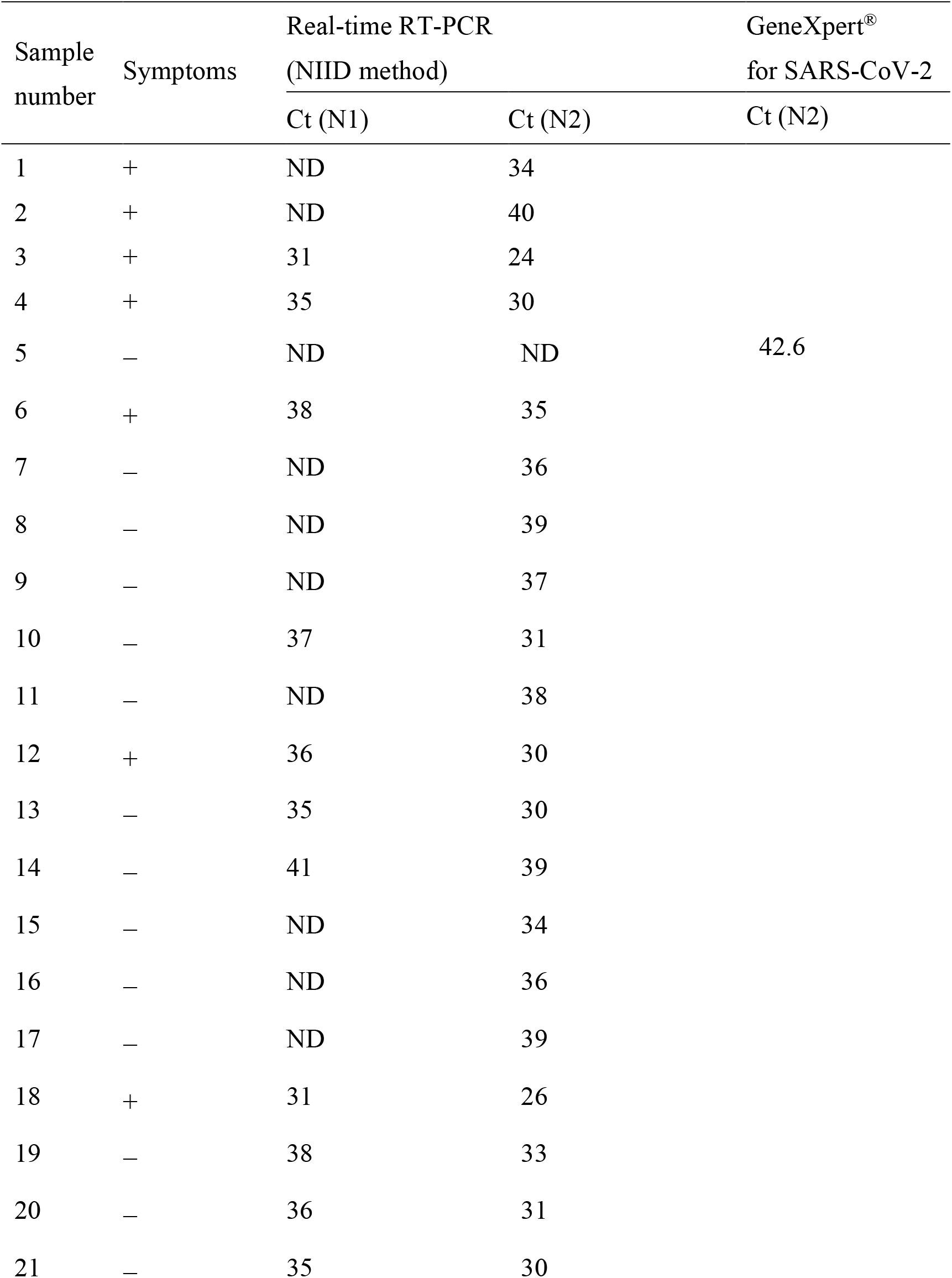
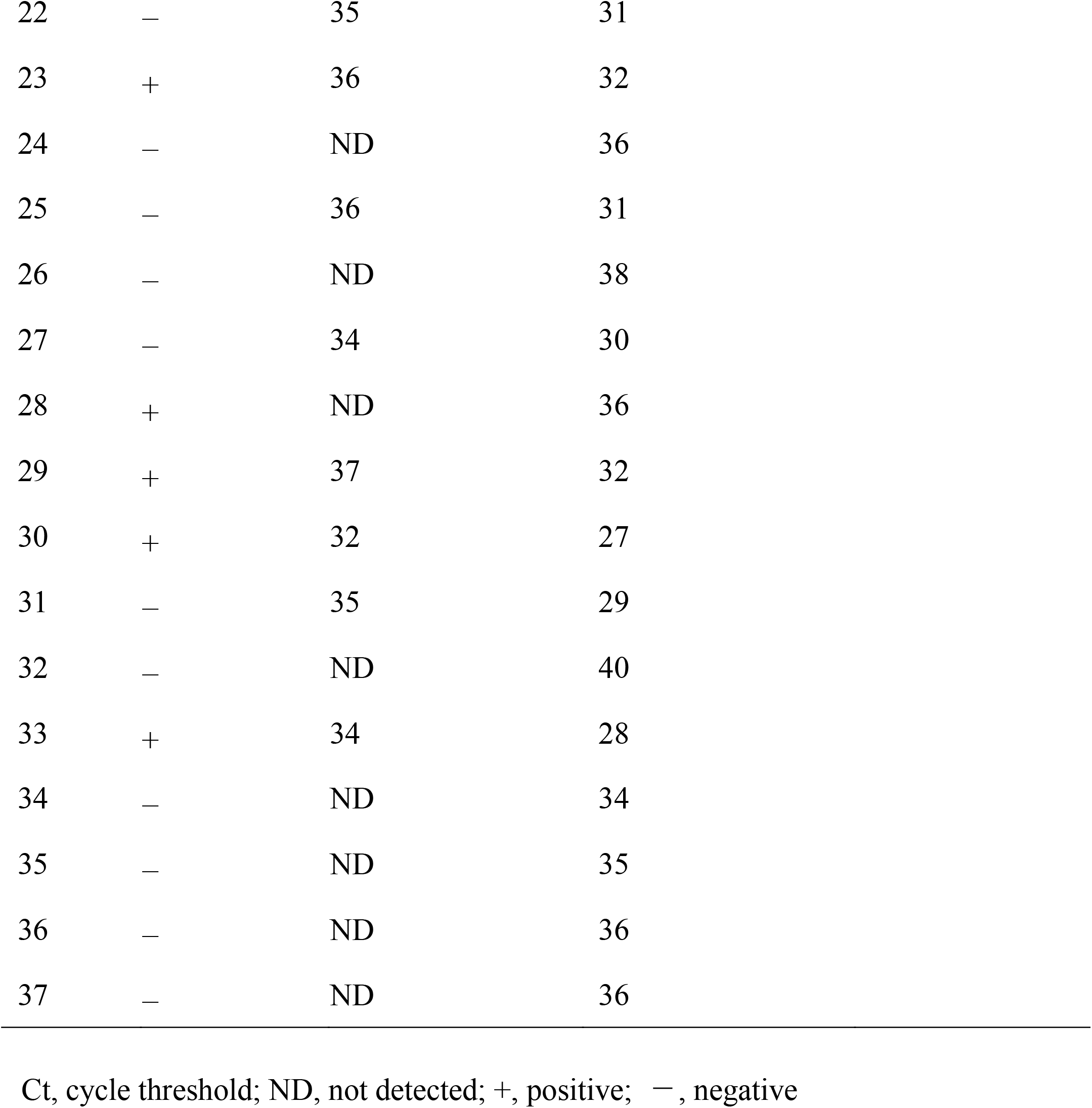
Detailed data of the 37 samples with discrepant findings between the antigen test and real-time RT-PCR.

| Sample number | Symptoms | Real-time RT-PCR<br>(NIID method) |  | GeneXpert®<br>for SARS-CoV-2 |
| --- | --- | --- | --- | --- |
|  |  | Ct (N1) | Ct (N2) | Ct (N2) |
| 1 | + | ND | 34 |  |
| 2 | + | ND | 40 |  |
| 3 | + | 31 | 24 |  |
| 4 | + | 35 | 30 |  |
| 5 | – | ND | ND | 42.6 |
| 6 | + | 38 | 35 |  |
| 7 | – | ND | 36 |  |
| 8 | – | ND | 39 |  |
| 9 | – | ND | 37 |  |
| 10 | – | 37 | 31 |  |
| 11 | – | ND | 38 |  |
| 12 | + | 36 | 30 |  |
| 13 | – | 35 | 30 |  |
| 14 | – | 41 | 39 |  |
| 15 | – | ND | 34 |  |
| 16 | – | ND | 36 |  |
| 17 | – | ND | 39 |  |
| 18 | + | 31 | 26 |  |
| 19 | – | 38 | 33 |  |
| 20 | – | 36 | 31 |  |
| 21 | – | 35 | 30 |  |

|  |  |  |  |
| --- | --- | --- | --- |
| 22 | — | 35 | 31 |
| 23 | + | 36 | 32 |
| 24 | — | ND | 36 |
| 25 | — | 36 | 31 |
| 26 | — | ND | 38 |
| 27 | — | 34 | 30 |
| 28 | + | ND | 36 |
| 29 | + | 37 | 32 |
| 30 | + | 32 | 27 |
| 31 | — | 35 | 29 |
| 32 | — | ND | 40 |
| 33 | + | 34 | 28 |
| 34 | — | ND | 34 |
| 35 | — | ND | 35 |
| 36 | — | ND | 36 |
| 37 | — | ND | 36 |

Ct, cycle threshold; ND, not detected; +, positive; —, negative

### Comparison of the Ct values of samples from symptomatic and asymptomatic subjects

The median Ct values for the N2 gene of the samples from symptomatic and asymptomatic volunteers were 20 and 24, respectively. The N2 gene Ct values for samples from asymptomatic individuals were significantly higher than those for samples from symptomatic patients (*p* < 0.001; Fig. 1). The relationship between Ct values and days post-symptom onset is shown in Supplementary Figure 1.

**Figure 1.**
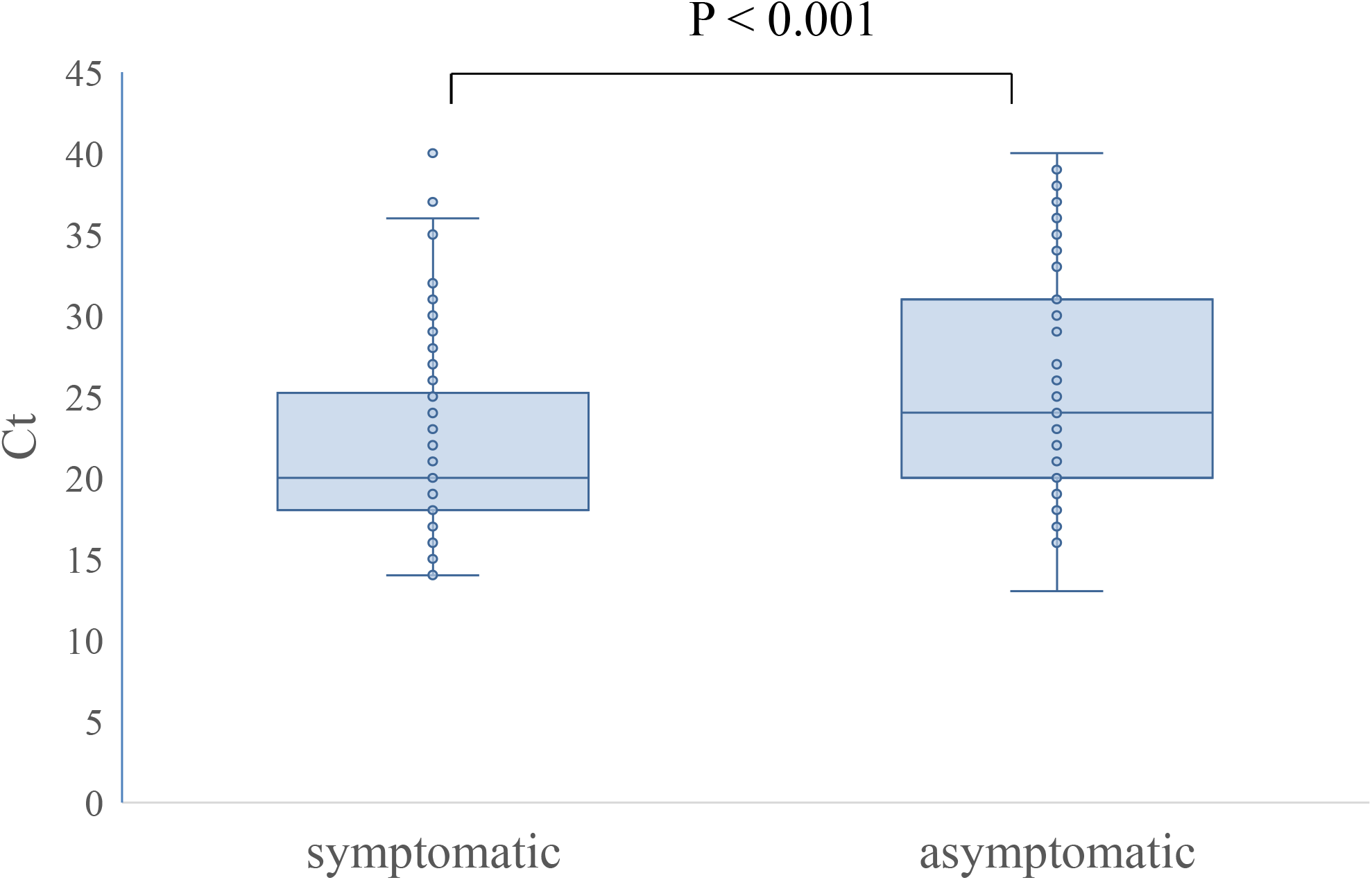
Ct values of samples from symptomatic and asymptomatic subjects. The number next to each box indicates the median Ct value.

## Discussion

Among the 1,073 samples collected from asymptomatic volunteers, the QuickNavi™-COVID19 Ag showed a sensitivity of 67.1% (95% CI: 55.4%–77.5%) and specificity of 100% (95% CI: 99.4%–100.0%). The sensitivity of this test for asymptomatic individuals was significantly lower than its sensitivity for symptomatic patients. No false positives were identified on the antigen test results among all 1,934 samples.

Studies of SARS-CoV-2 outbreaks have found that asymptomatic individuals comprise a significant portion of the infected population. [8]. A study of the outbreak on the Japanese cruise ship *Diamond Princess* reported that 328 out of the 634 confirmed SARS-CoV-2-infected patients were asymptomatic at the time of diagnosis [9]. It has been suggested that infections in asymptomatic individuals play an essential role in the SARS-CoV-2 epidemic [10], and efficient detection of asymptomatic individuals is necessary to control future outbreaks [11]. Because a symptom-based SARS-CoV-2 screening approach is incapable of detecting infected individuals who are asymptomatic, large-scale testing is needed for successful contact tracing. However, it is challenging to perform NAATs on a large scale and at high frequency owing to the limited capacity of laboratories and testing supplies [12]. In this respect, the simplicity and rapidity of antigen testing may make it useful for large-scale COVID-19 screening.

There is limited data on the performance of antigen tests in asymptomatic SARS-CoV-2-infected individuals [13]. The current study found that the sensitivity of our antigen test was lower in asymptomatic individuals than in symptomatic patients (67.1% vs. 89.3%). The median viral load may be lower in persons lacking symptoms (Fig. 1), which could explain this finding [14]. A few studies have observed a similar trend of lower antigen test sensitivity in asymptomatic individuals (45.4% vs. 79.1% [15], 53.3% vs. 84.6% [14]). Nevertheless, the lower sensitivity of COVID-19 antigen tests for asymptomatic individuals does not necessarily negate the utility of these assays for screening purposes. A model study showed that the high frequency and short sample-to-result time of tests might be more important than test accuracy for controlling COVID-19 outbreaks via population screening [16]. Furthermore, large-scale screening for SARS-CoV-2 infection demands high specificity to avoid unnecessary further examinations or the application of quarantine measures, which also have social costs [4]. We propose that the QuickNavi™-COVID19 Ag is a reasonable candidate for use in COVID-19 screening tests, given its very high specificity and lack of false-positive results in 2,796 samples (1,934 nasopharyngeal samples in the present study and 862 anterior nasal samples in our previous study [17]).

Among the group of samples with Ct values of <30, the QuickNavi™-COVID19 Ag showed a sensitivity of >80% for both symptomatic and asymptomatic subjects. This sensitivity met the performance requirement by the World Health Organization, which suggests that a sensitivity of ≥80% is “acceptable” in samples with Ct values of 25–30 [18]. Of the 37 false-negative samples, the majority had a low viral concentration, and only five had Ct values of <30. As indicated in a previous report, patients with low viral shedding have low infectivity [19]. Thus, the risk of overlooking high infectivity patients with COVID-19 in cases when the antigen test provides negative results seems limited. However, asymptomatic individuals who are in the early phase of SARS-CoV-2 infection may later develop symptoms with progressively increasing viral shedding [8]. Therefore, in addition to conducting COVID-19 screenings, symptom follow-up in asymptomatic individuals is essential.

Several limitations associated with the present study warrant mention. First, the results were obtained at a single PCR center during one epidemic season. Whether the same results would be obtained in other regions or during other epidemics requires additional validation. Second, this study did not include patients who had received the vaccine, and the accuracy of the test on SARS-CoV-2-infected patients after vaccination needs to be verified in the future. Third, we did not perform a genetic analysis of the detected SARS-CoV-2 variants and did not study the effect of genetic mutation on the antigen test results. Nevertheless, according to manufacturer’s information for use (version 4.0), QuickNavi™-COVID19 Ag reacts with both the SARS-CoV-2 UK variant (VOC-202012/01) and the Brazilian variant (501Y.V3, P.1), and the degrees of reaction with these variants are the same as those with the Wuhan strain.

In conclusion, despite showing very high specificity, the QuickNavi™-COVID19 Ag has lower sensitivity for detecting SARS-CoV-2 in asymptomatic individuals as compared with its sensitivity in symptomatic patients. Nevertheless, given its high convenience and specificity, this antigen test could be used as a supplementary COVID-19 assessment for asymptomatic individuals, as long as the results are interpreted appropriately.

## Data Availability

The data that support the findings of this study are available from the corresponding author, Y. K., upon reasonable request.

## Acknowledgments

We thank Mrs. Yoko Ueda, Mrs. Mio Matsumoto, Dr. Yumi Hirose, and the staff in the Department of Clinical Laboratory of Tsukuba Medical Center Hospital for their intensive support of this study. We thank all of the medical institutions for providing their patients’ clinical information. Mrs. Yoko Ueda and Mrs. Mio Matsumoto significantly contributed to creating the database for this study.

## Figure Legends

Supplementary Figure 1. Ct values and days post-symptom onset. Blue and red dots indicate true-positive and false-negative QuickNavi™-COVID19 Ag results, respectively.

