## Supplementary figures and images for "Prospective analytical performance evaluation of the QuickNavi™-COVID19 Ag for asymptomatic individuals"

### supplementary figure

#### Supplementary Figure 1

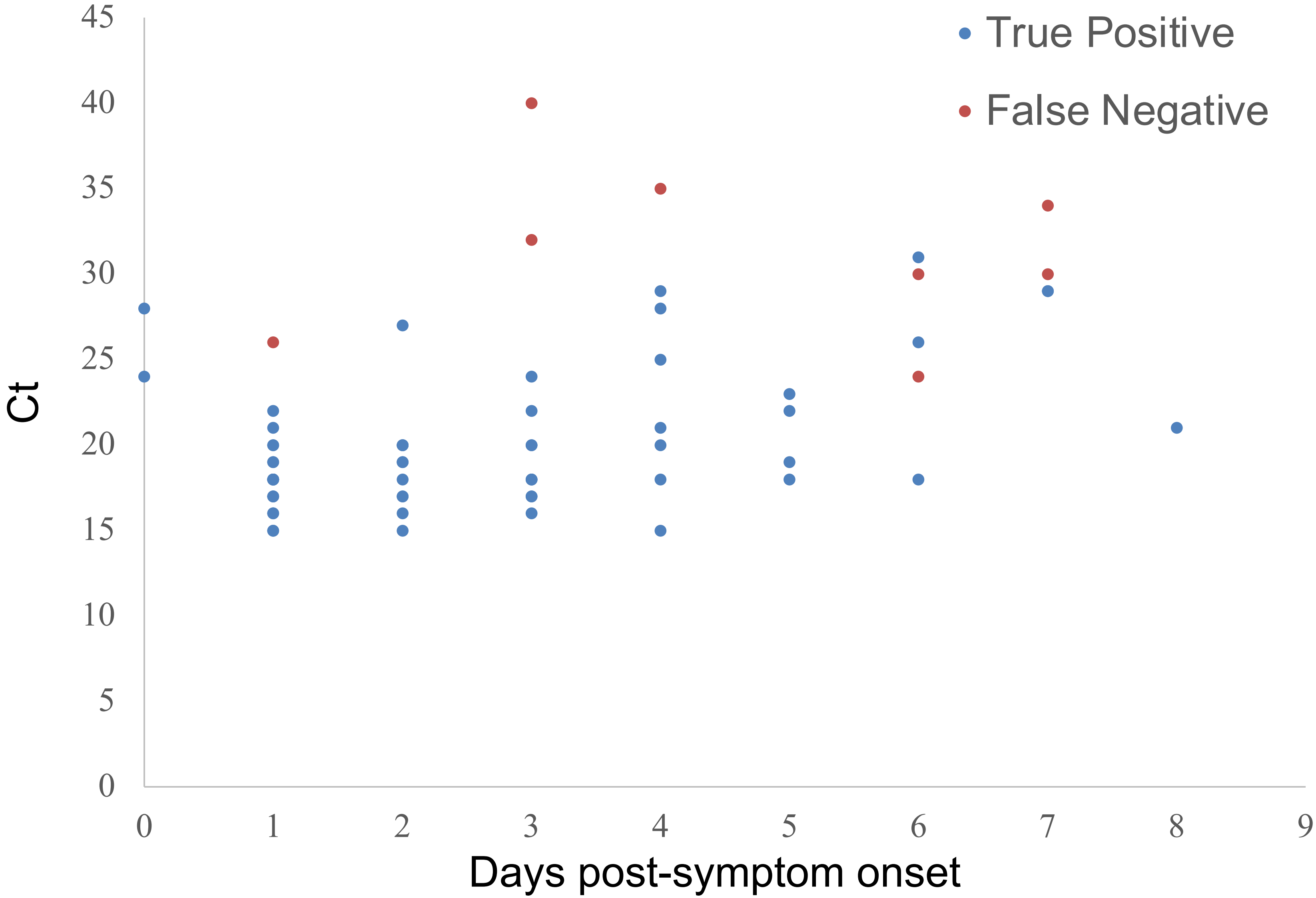
